# Simultaneous co-circulation of two genotypes of dengue virus serotype 3 causing a large outbreak in Sri Lanka in year 2023

**DOI:** 10.1101/2024.05.09.24307112

**Authors:** Dinuka Ariyaratne, Bhagya Senadheera, Heshan Kuruppu, Tibutius Thanesh Pramanayagam Jayadas, Laksiri Gomes, Diyanath Ranasinghe, Farha Bary, Ananda Wijewickrama, Sully Márquez Aguilar, Shannon Bennett, Chandima Jeewandara, Gathsaurie Neelika Malavige

## Abstract

As many other countries, Sri Lanka experienced a marked rise in the number of dengue cases in 2023, with an unusual pattern of disease epidemiology. This rise coincided with the emergence of dengue virus (DENV) serotype 3 in Sri Lanka as the predominant serotype after 2009. Interestingly, a discrepancy between NS1 rapid antigen test positivity and quantitative real time PCR positivity was observed, with 50% of NS1 positive samples being negative by molecular diagnostics. Following sequencing of the DENV-3 strains in 2023, we identified two DENV-3 genotypes (I and III) co-circulating. While DENV-3 genotype III was detected by the modified CDC DENV-3 primers, genotype I evaded detection due to key mutations at forward and reverse primer binding sites. The co-circulation of multiple genotypes associated with an increase in cases highlights the importance of continuous surveillance of DENVs to identify mutations resulting in non-detection by diagnostics and differences in virulence.

**One-sentence summary line:** Co-circulation of two genotypes of the dengue virus serotype-3 (I and III) were responsible for the large dengue outbreak in Sri Lanka in 2023, with genotype I not been detected by certain PCR primers.

## Introduction

Dengue is one of the most rapidly emerging mosquito-borne viral infections globally, with significant increases in morbidity and mortality (1). An emergency health warning was issued by WHO in 2024 due to the exponential rise in cases (2). The WHO named dengue as one of the top ten threats to global health in 2019 (3). However, during the COVID-19 pandemic, many countries observed a decrease in dengue incidence, reporting low to moderate levels of transmission (4). With the cessation of the global health emergency for COVID-19, the incidence of dengue has markedly increased, attributed to climate change, fragile health systems, political, economic and humanitarian crises, unplanned urbanization and population movement (5, 6).

Sri Lanka has experienced regular outbreaks of dengue since 1989, with the number of cases gradually increasing over the years and spreading into different geographical locations within the country (7). The largest outbreak due to dengue was reported in 2017 with 186,101 cases, associated with the cosmopolitan strain of the dengue virus (DENV) serotype 2 (4, 8). DENV-2 continued to be the predominant circulating serotype until October 2019. DENV-3 began to emerge towards the end of October, with an increase in the number of dengue cases, with a total of 105,049 cases being reported (4). 20,718 cases (one fifth) were reported from Colombo, which is usually the district with the highest incidence. However, with the onset of the COVID-19 pandemic, the number of dengue cases decreased drastically in 2020 (4257 reported in Colombo) and in 2021 (11,401 reported in Colombo) (9). During these two years, DENV-2 accounted for the majority of cases (46%), followed by DENV-3 (27.9%) and DENV-1 (24.1%) ((4). In Sri Lanka, the number of cases began to gradually increase from June 2022 onwards with a total number of 89,799 cases reported in 2023, with 18,650 from Colombo (10). Usually, Sri Lanka has two seasons of intensified dengue activity coinciding with the monsoon seasons (7, 10). One season typically spans November to early February and the second season runs from May to July. However, 2023 saw an unusual pattern, since outbreak-level transmission persisted from November 2022 until July 2023 without inter-monsoon respite.

As we have been carrying out dengue surveillance activities in Colombo, Sri Lanka for many years, we tracked these dynamics and noted an unusual discrepancy between NS1 antigen test results and real-time PCR results for serum samples from patients with a suspected acute dengue infection. Our surveillance activities shows that the outbreak coincided with the emergence of DENV-3 serotype as the predominant circulating serotype in Colombo. The aim of this study was to understand the unusual DENV-3 outbreak pattern as well as the discrepancy between point of care diagnostics and molecular diagnostics. To this end, we sequenced virus-positive samples to identify the serotype of infecting DENVs along with other genetic information.

## Methods

### Patients

From June 2022 to November 2023, we tested serum samples of 210 patients with a suspected acute dengue infection admitted to the National Institute of Infectious Diseases, Colombo, following informed written consent. All samples were collected from patients presenting with symptoms suggestive of an acute dengue infection, within the first five days since onset of illness. The NS1 rapid antigen test (SD Biosensor, South Korea) was conducted for a randomly selected sub-cohort of patients at the time of recruitment. The samples were transported to our laboratory and serum was separated and stored at −80 C.

### Ethics approval

Ethical approval was obtained from the Ethics Review Committee of the Faculty of Medical Sciences, University of Sri Jayewardenepura.

### Quantitative real-time PCR assays for detection of the DENV serotypes

Viral RNA was extracted from serum samples using MagMAX™ viral nucleic acid isolation kit (Applied Biosystems, USA, Cat: A48310) and an automated KingFisher™ Flex purification system (Applied Biosystems, USA). Quantitative real time PCR was performed using oligonucleotide primers published by the CDC with slight modifications as described previously (4, 11). A dual labelled probe for detection of DENV serotypes 1 to 4 was used (Life technologies, USA) based on published sequences (Life Technologies, India) (4). However, in 2023, 50% of the NS1 antigen positive samples (Table 1) gave a negative result with the qRT-PCR protocol used by us since 2014, we also implemented the Realstar Dengue Type RT-PCR Kit 1.0 (Altona diagnostics, Germany), in a subset of NS1 positive, but CDC PCR protocol negative samples (n=50) according to manufacturer’s instructions.

**Table 1:** The PCR and NS1 antigen positivity rates from 2017 to 2023 tested in our laboratory. The number of acute dengue patients’ samples from Colombo that were tested using NS1 rapid antigen test and PCR using CDC primers are shown, along with the percentage positivity rates of both tests compared to the number of patients clinically diagnosed and managed as dengue.

| Year | Number of samples tested for PCR and positivity rates<br>Positive<br>N (%) | Number of samples that were tested for NS1 and positivity rates | Number of NS1 positive samples that were negative for PCR |
| --- | --- | --- | --- |
| 2017 | 26/32 (81.25%) | 31/31 (100%) | 5/31(16.1%) |
| 2018 | 162/169 (95.85%) | 85/169 (50.2%) | 3/85 (3.5%) |
| 2019 | 233/254 (91.73%) | 244/253 (96.4%) | 20/244 (8.1%) |
| 2022 | 81/104 (77.8%) | 53/66 (80.3%) | 10/53 (18.9%) |
| 2023 | 68/106 (43%) | 136/179 (75.9%) | 68/136 (50%) |

### Library Preparation and Next Generation Sequencing (NGS) of Dengue 3

Extracted viral RNA (QIAamp viral RNA mini kit, Qiagen, United States) was quantified (NanoDrop™ 2000c Spectrophotometers, Thermo Fisher Scientific, USA) and then reverse transcribed using LunaScript™ RT SuperMix. Tiling PCR was then performed using specific dengue primers for DENV-3 as previously published (12). Briefly, two primer pools (10µM) A and B were prepared, each mixed with Q5 DNA polymerase (Oxford Nanopore Technologies) and nuclease free water. After preparation, the mixtures were amplified by denaturing them at 98°C for 30 seconds, followed by 45 cycles of 98°C for 15 seconds and 65°C for 5 minutes. Lastly, they were held at 4°C using a QuantStudio™ 5 Real-Time PCR Instrument (Applied Biosystems, Singapore). Amplicons of 900bp (12) were verified by gel electrophoresis. Subsequently, rapid barcode ligation was performed using Rapid Barcoding kit (SQK-RBK110.96, Oxford Nanopore Technologies). Barcoded amplicons were pooled and cleaned up with 70% ethanol using SPRI beads (Oxford Nanopore Technologies). The libraries were then quantified using the Qubit dsDNA High Sensitivity assay kit on a Qubit 4.0 instrument (Life Technologies) and 1000ng of libraries in 11 μl with the addition of Rapid Adapter-F loaded on to a MinION flow cell (FLO-MIN 106, Oxford Nanopore Technologies) and run on a Mk1C (Oxford Nanopore Technologies). Basecalling was performed using Guppy (version 6.5.7) with Fast model, 450 bps basecalling model. Reads were aligned with DENV-3 reference genome (NCBI accession no. NC_001475.2 Dengue virus 3) using the wf-alignment workflow. Next, Samtools (Version 1.16.1) was used to construct consensus sequences (Minimum quality score threshold per base: 20 and Minimum depth coverage per base for consensus: 10). Genotype assignment was performed with the Flavivirus Genotyping Tool Version 0.1.

### DENV-3 time-resolved phylogenetic tree

We subjected six whole genome sequences with >70% coverage of the DENV-3 genome (NC_001475.2) and >90% coverage of the E and NS1 regions to phylogenetic analysis. Accession numbers included in the analysis are as follows: SRA PRJNA1108507 (genome which has full coverage. PP767447, PP767459, PP766873, PP770477, PP770484, PP770483 (genomes which have complete coverage of NS1 and regions). To carry out phylogenetic analysis, a total of 1316 DENV-3 complete genomes were downloaded from the Genbank database. The matching of the whole genomes with the corresponding metadata was perform using with R\Biostrings (v.2.70.1) and R\BSgenome packages (v.1.70.1) run on R version 4.3.2. The extracted 1316 samples including 6 samples sequenced by our laboratory in 2023 (named as “Sri Lanka 2023” in the phylogenetic trees), were used for the multiple sequence alignment. Multiple sequence alignment was generated using MAFFT v.7.508 employing the FFT-NS-i algorithm.

Subsequently, this multiple sequence alignment was used to infer a Randomized Axelerated Maximum Likelihood (RAxML) phylogenetic tree using RAxML (v.8.2.12) with GTRGAMMA substitution model and bootstrap of 1000 replicates (13). The best-fit model GTR+F+R5 was chosen using ModelFinder (14). Subsequently, a molecular clock phylogeny was generated using Clockor2 (v.1.6.6). Final visualizations of the phylogenetic tree were done using R\ggtree, R\ape and R\ggstar packages (R version 4.1.2).

Molecular clock phylogenetic analysis was carried out with these 1316 DENV-3 sequences extracted from Genbank originating from 46 countries, from years 1956 to 2023, by using the Maximum Likelihood-based TreeTime version 0.10.1 [7]. Two outliers were detected and removed during the root-to-tip regression analysis (substitution rate: 7.864e-04, R^2^: 0.78). Further, maximum local trees were analyzed with Best-Fit-Model. Final visualizations of the phylogenetic tree were done using R\ggtree, R\ape and R\ggstar packages (R version 4.1.2).

### Mutation Analysis

We performed a mutation analysis of the primer and probe binding regions of the CDC primers, located between nucleotides 740-813, which correspond to the nucleotides coding the membrane region of the DENV-3 genome (Supplementary Figure 2). We used the multiple sequence alignment to detect the mutations within the DENV-3 strains identifies in Sri Lanka in 2013, in comparison to sequences from previous years in Sri Lanka and globally. We incorporated the mutation patterns in the primer binding regions and included a minimum of five level of ancestral branches of our samples in the phylogenetic tree constructed in the previous analysis. Sequence visualizations of the phylogenetic tree were generated using R\ggtree, R\ggplot and R\msa packages (R version 4.1.2).

## Results

### Serotype detection

We have conducted DENV surveillance activities at the National Institute of Infectious Diseases, Colombo since 2014, which is one of the largest infectious disease hospitals in Sri Lanka. Until the later part of 2022, the proportion of dengue NS1 positive samples, which were negative by PCR remained below 16.1% (Table 1). The rates of samples positive for NS1 antigen but negative by PCR were 18.9% (10/53) in 2022 and rose to 50% (68/136) by November 2023 (Table 1). Using the Realstar Dengue Type RT-PCR Kit 1.0 (Altona diagnostics, Germany), the DENV serotype could be identified in 31/50 of the samples previously negative according to the CDC primer-based RT-PCR. From these, 15/50 samples were identified as DENV-3, 14 samples as DENV-2 and 2 samples as DENV-1.

### Phylogenetic analysis of the DENV-3 sequences in Sri Lanka

Sequencing was carried out in a total of 22 samples. 12 of these samples were CDC primer PCR positives and 10 were PCR negative using CDC primers. From these, only fourteen had adequate coverage and were assigned to their corresponding genotype. We found that 11 samples positive by the CDC DENV-3 primers were of DENV-3 genotype III, whereas the DENV-3 sequences that were not detected by the CDC DENV-3 primers corresponded to genotype I (n=3). Six of these sample genomes had a coverage greater than 70% of the whole DENV-3 genome and >90% coverage of the E and NS1 regions. These genomes were included in the phylogenetic analysis (Figure 1).

**Figure 1:**
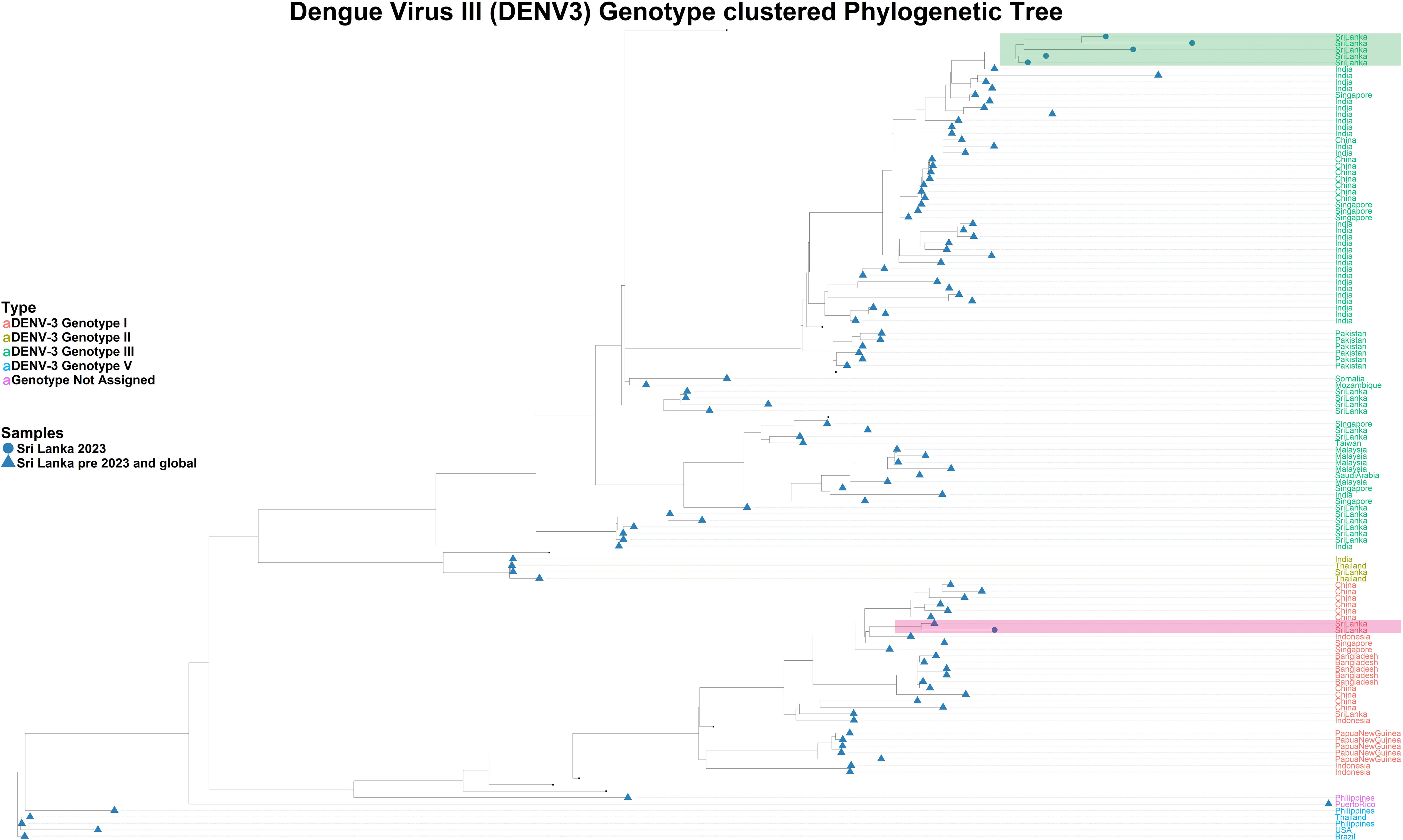
Phylogenetic tree of the DENV-3 viruses sequenced from Sri Lanka in 2023 in comparison to global DENV-3 sequences. The Sri Lankan DENV-3 viruses sequenced were analysed with 1316 sequences from 46 countries from 1956 to 2023. The Sri Lankan DENV-3 sequences, which were not detected by the CDC DENV-3 primers were assigned to genotype I (highlighted in green) and the DENV-3 sequences that were detected by CDC primers assigned to genotype III (highlighted in pink).

Three main clusters (Clade I, Clade II, Clade III) were identified in the phylogenetic tree of DENV-3 genotype I (Figure 2). Three sub clusters are identified within Clade I, which includes sequences from China, Sri Lanka, Bangladesh and Papua New Guinea. Genotype I of DENV-3 previously reported from Sri Lanka in 2017 was found to be closely related to the current 2023 strains of genotype I from Sri Lanka (Figure 2). The Sri Lankan DENV-3 genotype I sequences from 2017 and 2023, share a common ancestor with DENV-3 sequences from China (Clade I).

**Figure 2:**
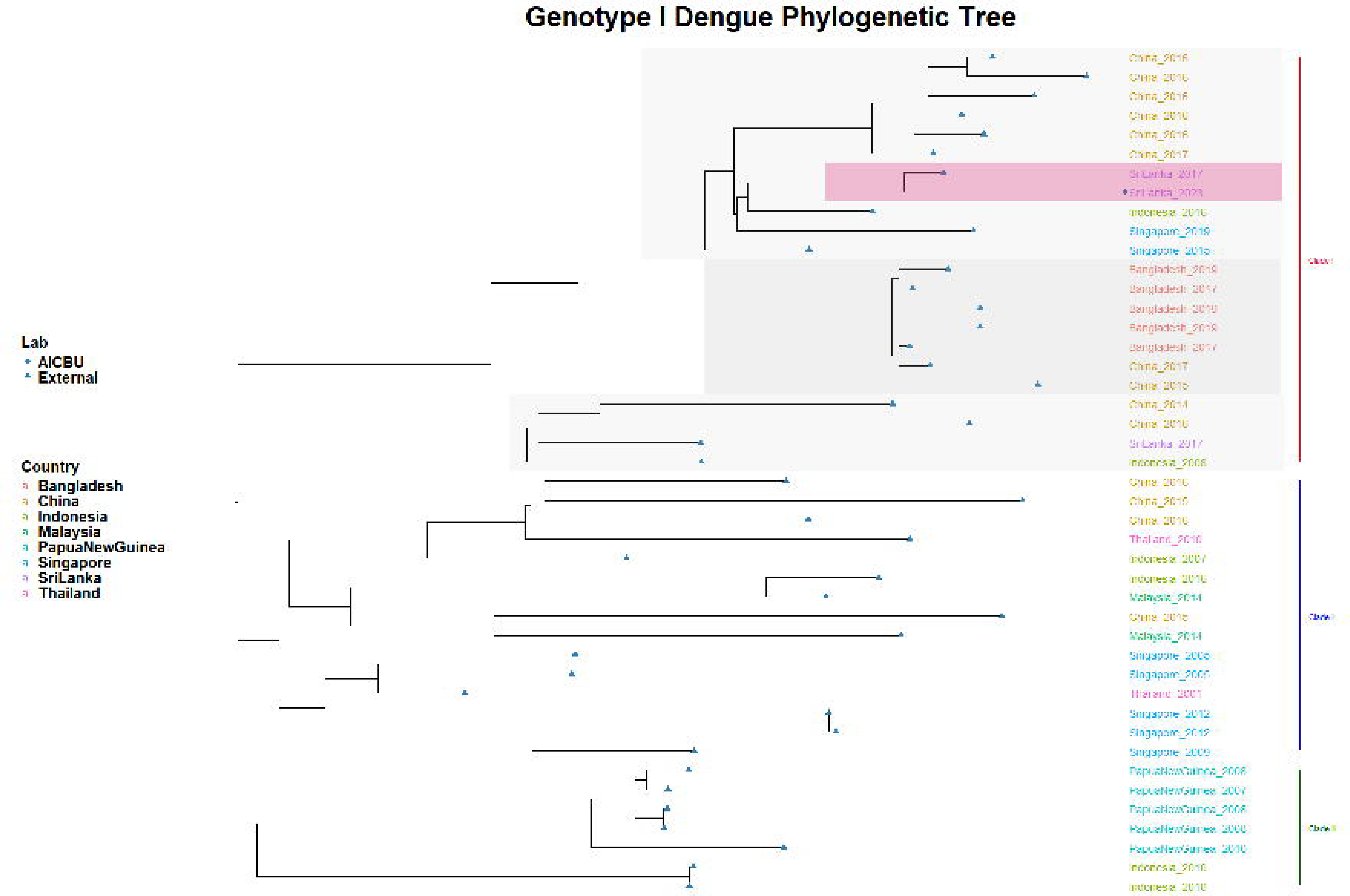
The phylogenetic tree of genotype I DENV-3 sequences. The phylogenetic tree was generated with the Sri Lankan genotype I strains in comparison to the global genotype I strains. Three main clusters (Clade I, Clade II, Clade III) were identified in the phylogenetic tree of DENV-3 genotype I (Figure 2). Three sub clusters are identified within Clade I, which includes sequences from China, Sri Lanka, Bangladesh and Papua New Guinea. The Sri Lankan genotype I strain was assigned to the Southeast Asian sub-cluster of Clade I, along with sequences from China (shaded in grey).

The other co-circulating genotype in Sri Lanka in 2023, DENV-3 genotype III strains shared a common ancestor with a sequence from India collected in 2022 and shows that the Sri Lankan samples closely relate to global samples (Clade I) (Figure 3). Three sub-clusters were identified within Clade I, which included Indian sequences that were similar to the Sri Lankan sequences identified in 2023, a subcluster consisting of sequences from China and Singapore and a subcluster with Indian sequences alone. Clade II and Clade III in Figure 3, represents two different time zones in the evolution. Sri Lankan DENV-3 genotype III strains before year 2000 were assigned to Clade III.

**Figure 3:**
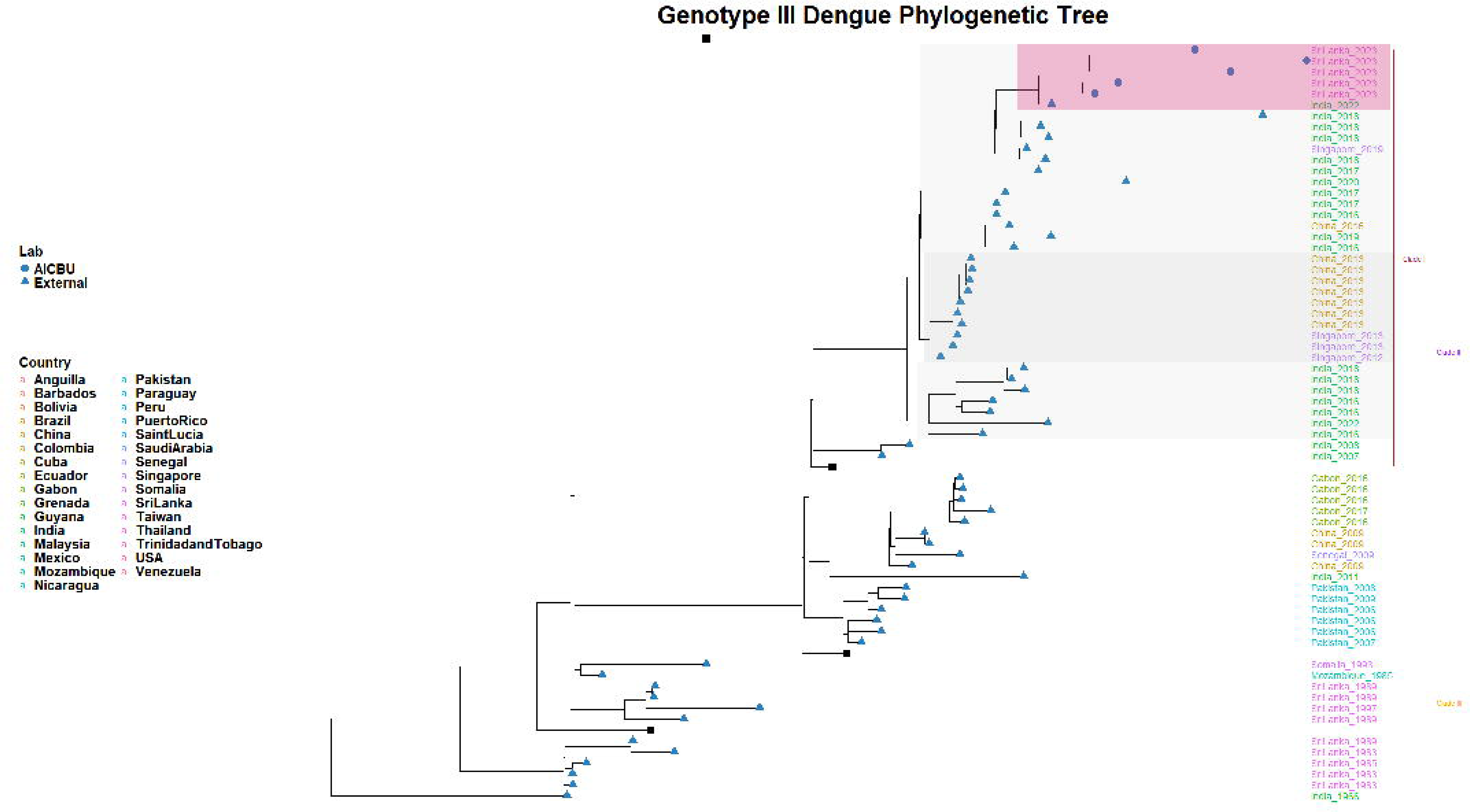
The phylogenetic tree of genotype III DENV3DENV-3 sequences. The phylogenetic tree was generated with the Sri Lankan genotype III strains in comparison to the global genotype III strains. All the Sri Lankan sequences from 2017 onwards were assigned to clade I, which consists of both South Asian and Southeast Asian DENV-3 genomes. Three sub-clusters were identified within Clade I, which included Indian sequences that were similar to the Sri Lankan sequences identified in 2023 (shaded in grey), a subcluster consisting of sequences from China and Singapore and a subcluster with Indian sequences alone.

### Molecular phylogenetic analysis

Molecular clock analysis was carried out to investigate the evolutionary rate of DENV-3 viruses using 1316 DENV-3 sequences from 46 countries, from 1956 to 2023. The evolutionary rate for DENV-3 was found to be 8.007 x 10^-4 nucleotide substitutions per site per year, with the root of the tree diverging into DENV-3 estimated to be year 1899 (124 years ago) (15). The molecular clock analysis for the Sri Lankan DENV-3 sequences found 21 nodes (for all genotypes), with an evolutionary rate of 7.950 x 10^-4 nucleotide substitutions per site per year (Figure 4). The molecular clock that mapped with genotype III showed an evolutionary rate of 7.409 x 10^-4 nucleotide substitutions per site per year. The second molecular clock mapped predominantly with genotype I, although a few other samples from other genotypes were also assigned to it. This second molecular clock (genotype I) showed an evolutionary rate of 6.814 x 10^-4 nucleotide substitutions per site per year. Our Sri Lankan DENV-3 strains demonstrated a high root to tip ratio, compared to the previous DENV-3 sequences, indicating a high mutation rate during the points of sampling (year 2017 to 2023).

**Figure 4:**
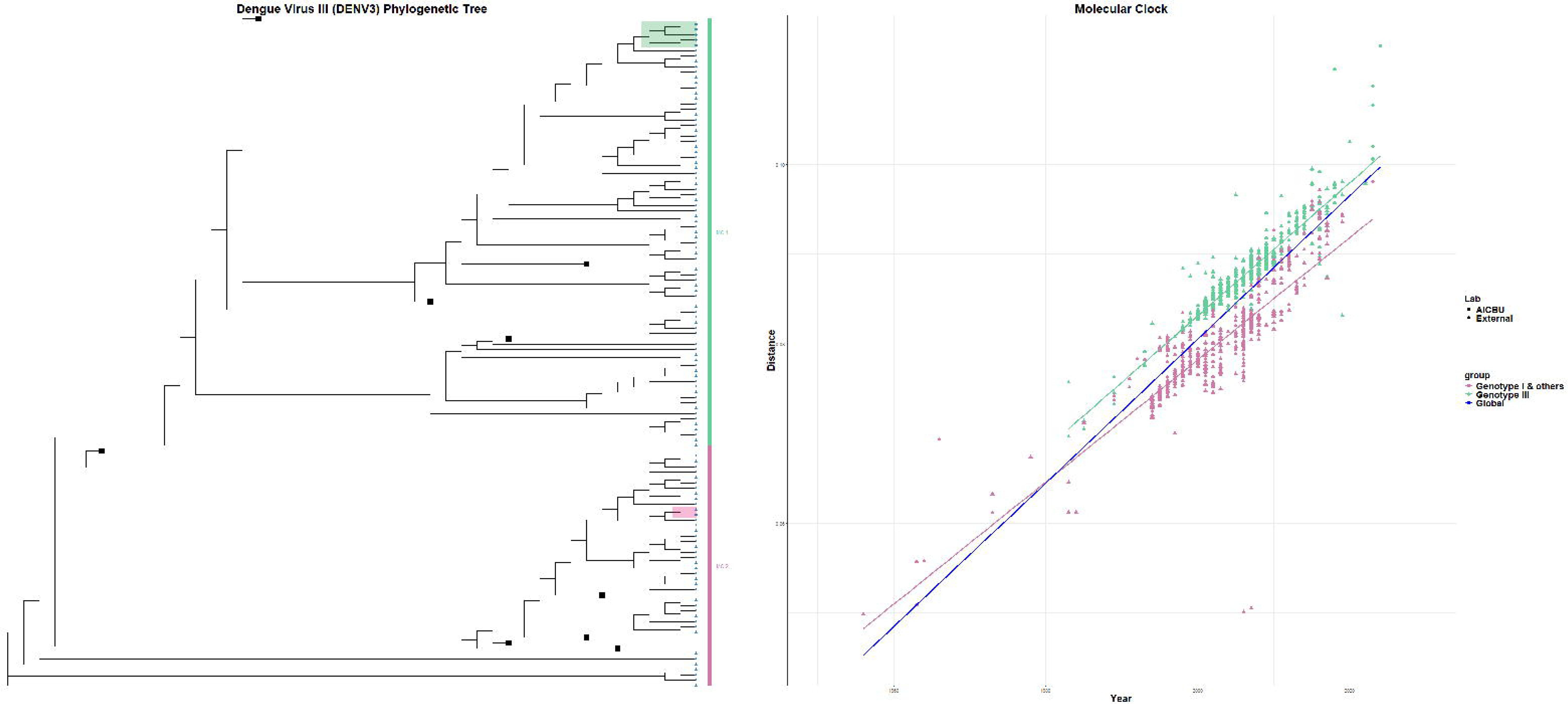
Evolution of the DENV-3 genotypes I and III. A molecular clock analysis (Root-to-Tip analysis) was carried out to investigate the evolution of DENV-3 genotypes I and III in Sri Lanka in relation to the global DENV-3 strains, which shows that both genotypes present in Sri Lanka co-evolved at a similar rate to the global dengue strains rate.

### Mutation analysis of primer binding regions

As the genotype I of DENV-3 was not detected by CDC DENV-3 primers, we proceeded to investigate the mutations that could be responsible for evading detection. We also investigated any mutations within the genotype III sequences of DENV-3. The regions of both genotype I and III were aligned with the forward and reverse primer binding sites and also the binding region of the probe (Supplementary figures 2 to 7). We found a single nucleotide substitution of C to T at position 744 in genotype I compared to the reference DENV-3 genotype I genome NC.001475.2. This mutation was located within the forward primer and reverse binding site of the CDC DENV-3 primers (Figure 5A and B) in Sri Lanka virus strains after 2017 and was also seen in the Sri Lankan genotype I, identified in 2023 (GenBank accession numbers: PP766873; SRA: PRJNA1108507 AICBU07.03_2023-06-05_SriLanka). These point mutations were detected within the Sri Lankan genotype I (2023) strain in positions C744T and A756G of the forward primer binding sites and in position G795A of the reverse primer binding sites. These mutations was not found in genotype III. The 740 to 820 region in genotype III was found to be more conserved compared to genotype I. There were no mutations detected within the probe binding region in genotype I or III (Figure 4C). DENV-3 strains with mutations within both the forward and reverse primer binding sites as seen with the DENV-3 genotype I strain was not detected elsewhere, based on published DENV-3 sequences.

**Figure 5:**
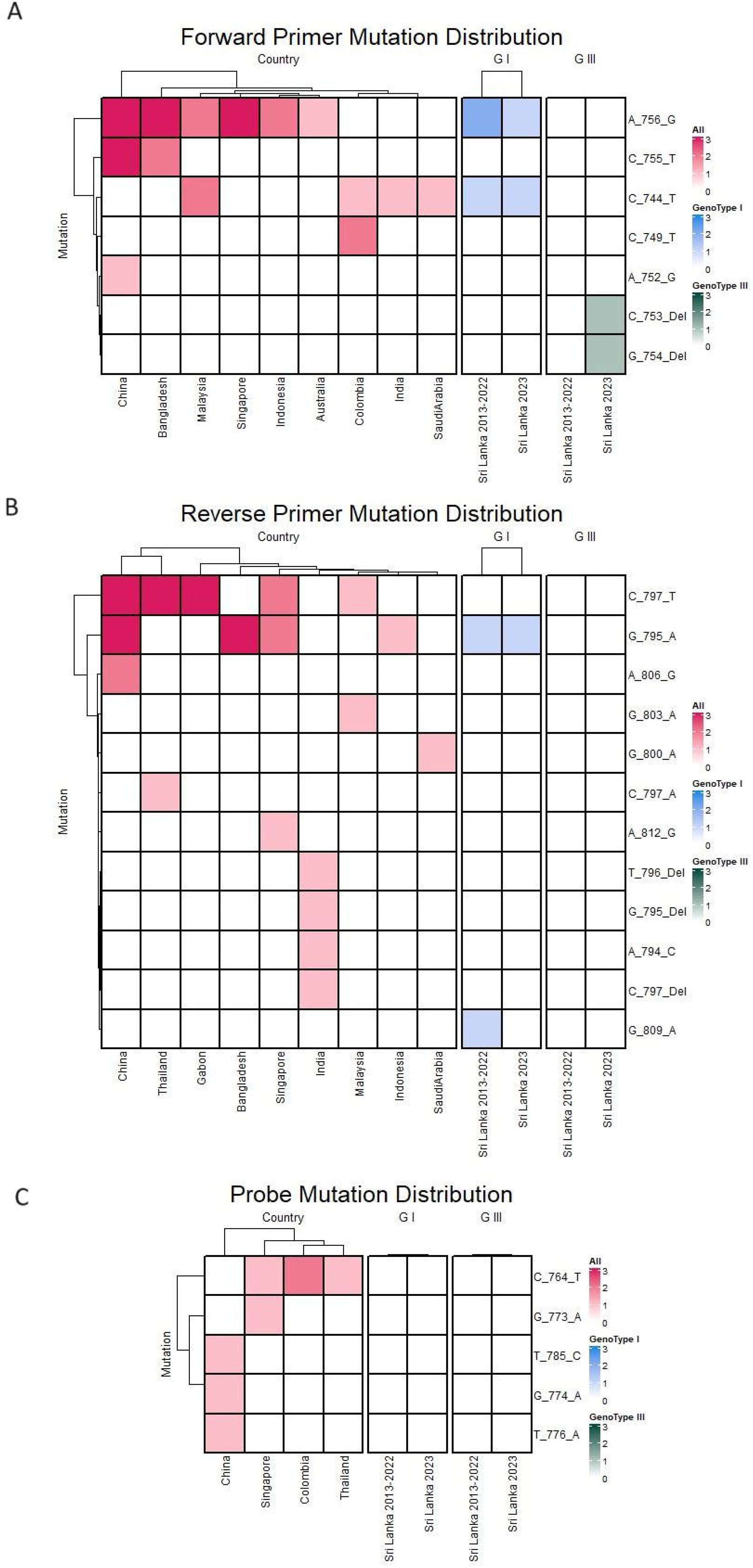
Mutation within the primer and probe binding regions for the CDC DENV-3 primers in the DENV-3 genotype I and III strains detected in Sri Lanka in 2023. The mutations within the CDC DENV-3 primer and probe binding regions (nucleotides 740-813) of Sri Lankan DENV-3 genotype I and III were compared with previous Sri Lankan DENV-3 strains and global DENV-3 strains reported between 2013-2023. Point mutations were detected within the Sri Lankan genotype I (2023) strain in positions C744T and A756G of the forward primer binding sites (nucleotide region 741-760), while no mutations were detected in the Sri Lankan genotype I (2023) strain (A). Point mutations were also detected within the Sri Lankan genotype I (2023) strain in position G795A of the reverse primer binding sites (nucleotide region 789-814), while no mutations were detected in the Sri Lankan genotype I (2023) strain (B), No mutations detected in either Genotype I or III in the probe binding site (nucleotide region 762-787).

## Discussion

The cosmopolitan strain of the DENV-2, which was responsible for the largest dengue outbreak contributed to be the predominant serotype until June 2022 (8). We detected a trend towards DENV-2 being replaced by DENV-3 starting June 2022, becoming the dominant serotype by 2023. We document the emergence of DENV-3 as the predominant serotype starting in 2019, likely responsible for the large outbreak reported in 2023. Interestingly, we found that two genotypes of the DENV-3 serotype (genotype I and III) were simultaneously co-circulating in Colombo, Sri Lanka. Although DENV-3 was seen to emerge by the end of 2019, due to the initiation of the COVID-19 pandemic, the dengue outbreak subsided, which was attributed to school closures and social distancing measures (4). When dengue re-emerged in mid-2022, DENV-3 appeared to have gradually replaced DENV-2. As the Sri Lankan population had not been exposed to DENV-3 for several years (8, 16), it is likely that the emergence of a ‘new’ serotype in a relatively naïve population gave rise to a large outbreak in 2023 with 89,799 cases.

During the DENV-3 outbreak, we observed a discrepancy between the NS1 antigen positivity rates and PCR positivity rates. The real-time PCR for dengue is more sensitive and specific compared to the NS1 antigen rapid test for diagnosis of acute dengue (17, 18). Based on our laboratory results prior to 2019, only 8.1% of samples that gave a positive result for the NS1 antigen test gave a negative result for PCR. However, in 2023, 50% of the samples that tested positive by NS1 rapid antigen test gave a negative result by PCR using the CDC primers. We noted that samples positive by NS1 antigen, yet PCR negative were more likely to be DENV-3 genotype I whereas those that were PCR positive were DENV-3 genotype III. NS1 antigen could persist longer than the DENV in patients with acute dengue and therefore, NS1 antigen detection methods such as ELISA could be more sensitive than PCR in detecting acute infection (19, 20). However, all samples tested in our study were during early illness (≤4 days of illness) and therefore, PCR has shown to have a higher sensitivity than detection of NS1. So far, such a discrepancy (50%) between PCR and NS1 results has not been reported elsewhere.

We identified point mutations, C744T and A756G of the forward primer binding sites and in position G795A of the reverse primer binding sites which were not identified in DENV-3 genotype III. In fact, DENV-3 strains with all three mutations were not identified elsewhere, based on published sequences. These critical mutations in the forward and reverse primer binding sites, simultaneously occurring in DENV-3 genotype I, are likely to have led to the PCR dropouts, which have not yet been reported elsewhere.

While one sequence of DENV-3 genotype I from Sri Lanka in 2017 has been recorded, DENV-3 was not detected in patients with acute dengue until the later part of 2019. Therefore, it is possible that DENV-3 genotype I was circulating in Sri Lanka, causing infection, but remained undetected by us and others due to PCR failure based on the primers used. Our results highlight the importance of systematic DENV surveillance including serotyping, genotyping and sequencing. Given the intense transmission seen today in most countries, accompanied by high rates of virus evolution, sequencing is critical to understand disease dynamics (6, 21, 22).

The genotype III of DENV-3 was shown to be responsible for the initiation of dengue outbreaks in Sri Lanka in the 1980s, which also caused dengue outbreaks globally (23). However, the current clade of the DENV-3 genotype III differs from strains circulating from 1980 to 1990 and is similar to those of the Indian subcontinent (24). The genotype III of DENV-3 causing outbreaks in Brazil in 2023 was also shown to originate from the Indian subcontinent via the Caribbean (25). Both genotypes I and III of DENV-3 have been responsible for recent outbreaks in Bangladesh, genotype III in India, and genotype I in Indonesia, although not so far being reported in many other Asian countries (24, 26, 27). DENV-3 genotype II has so far been responsible for most outbreaks in Southeast Asia (28).

Although many other countries experienced large dengue outbreaks globally and in South Asia due to DENV-3 (26), the genotypes and the clades responsible for these outbreaks in 2022 and 2023 have not been reported. Our molecular clock analysis showed that the genotype III of DENV-3 had been evolving at a faster rate than genotype I in Sri Lanka. Future studies should address the potential differences in phenotype of these two genotypes, for example differences in clinical disease severity and/or the vector competence. Furthermore, since many of the dengue vaccines incorporate only one genotype representing each of the DENV serotypes, it would be important to closely monitor the potential evolution of different genotypes to understand vaccine efficacy and possible immune evasion. As we have shown, characterizing sequence evolution is also critical for assessing the performance of molecular diagnostic tests.

## Data Availability

All data is available in the manuscript, figures and supporting files.

## Acknowledgement

We are grateful to the NIH, USA (grant number 5U01AI151788-02) for funding this study.

## Notes

### Competing Interest Statement

The authors have declared no competing interest.

