## Supplementary figures for "Simultaneous co-circulation of two genotypes of dengue virus serotype 3 causing a large outbreak in Sri Lanka in year 2023"

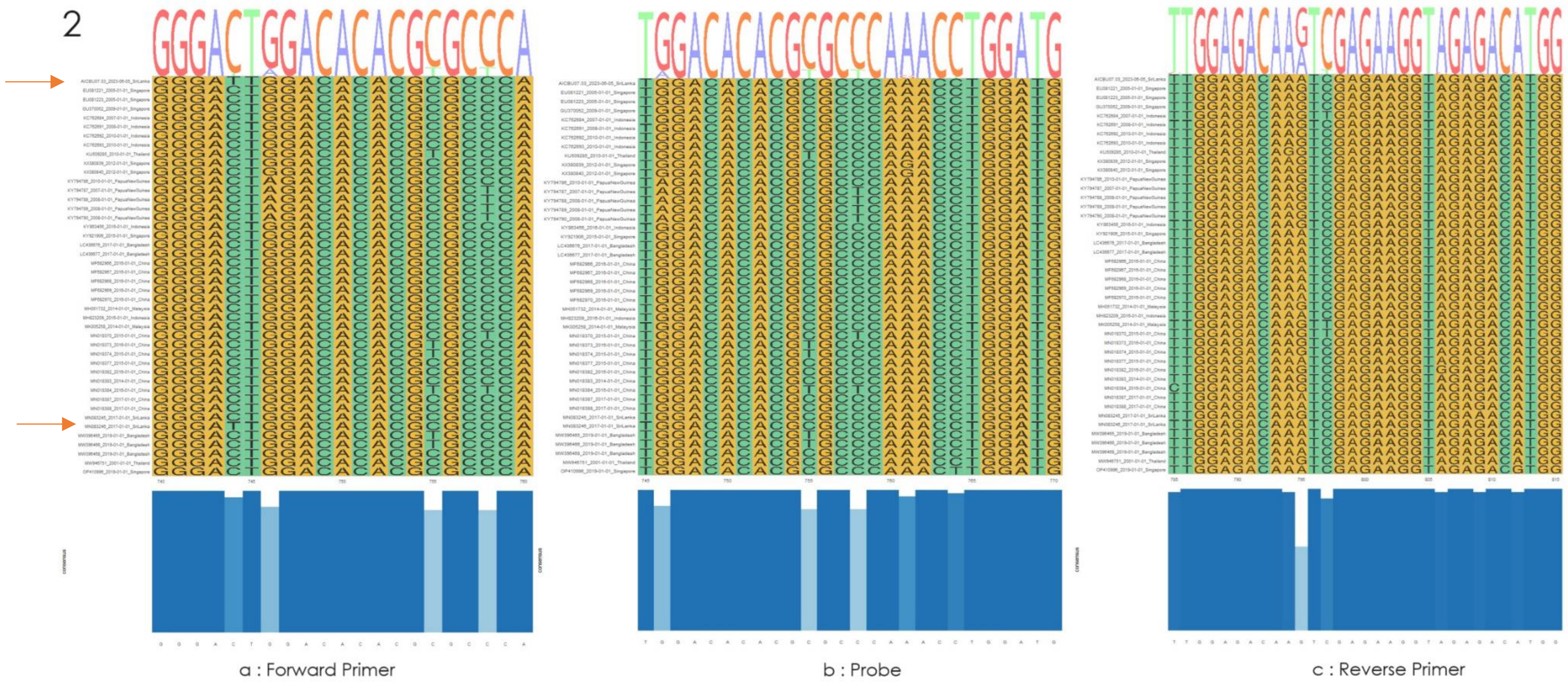


**Figure 1 :** Genotype I Primer and Probe binding regions in the M protein encoding area: a) The forward primer binding to nucleotides 740-750 the 2 Sri Lankan genotype I samples (red arrows) show a mutation C744T b) The probe binding to nucleotides 745-759 c)the reverse primer binding to nucleotides 788-813

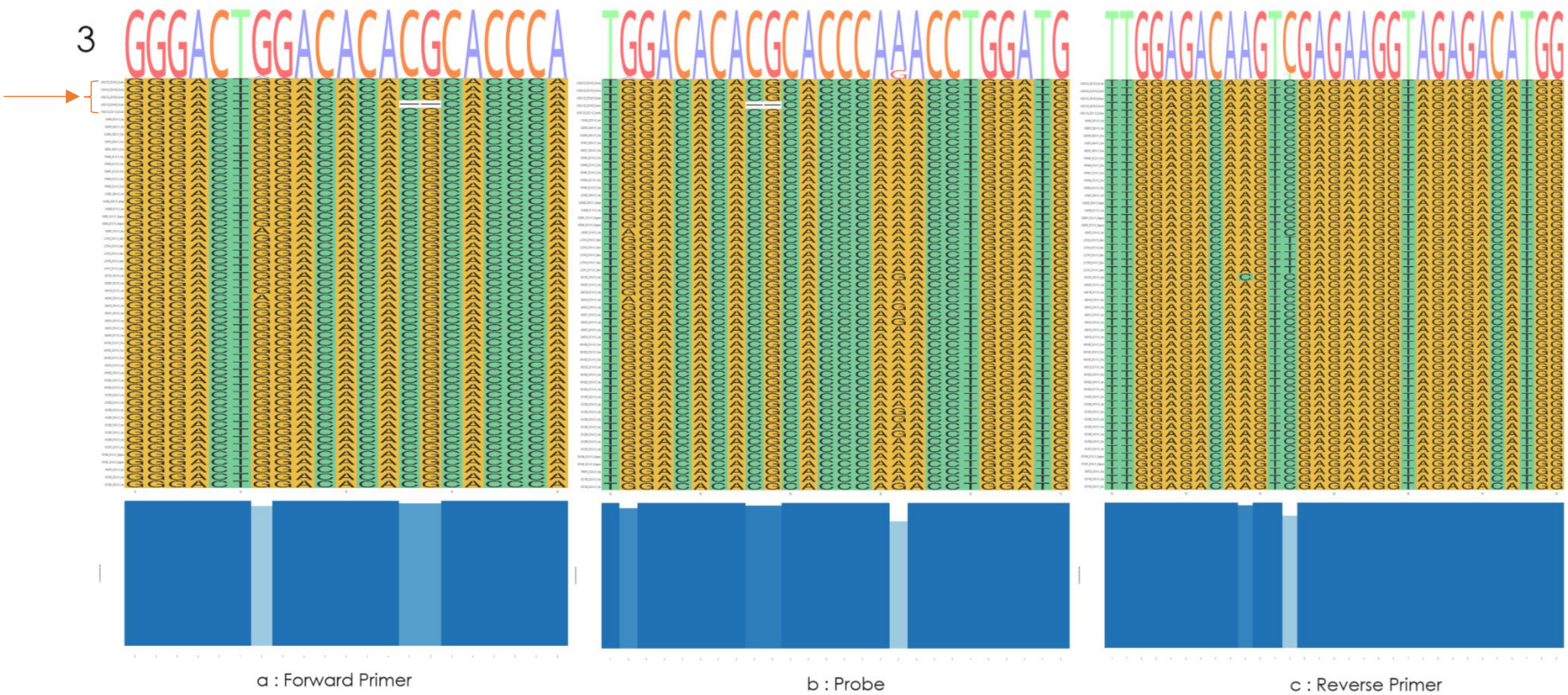


**Figure 2 :** Genotype III Primer and Probe binding regions in the M protein encoding area: a) The forward primer binding to nucleotides 740-750 the 5 Sri Lankan genotype I samples (red arrow & bracket) have no mutations in this region b) The probe binding to nucleotides 745-759 c) The reverse primer binding to nucleotides 788-813
